# PHGDH activity in Triple-Negative Breast Cancer studies--A glimmer of hope in the midst of difficult treatment process

**DOI:** 10.1101/2025.10.13.25337860

**Authors:** Yijun Xie, Ziyu Jiang, Zhiheng Huang, Zilong Chen, Hui Lyu, Ling He

## Abstract

**Background:** Triple-Negative Breast Cancer (TNBC), which lacks estrogen receptor, progesterone receptor and HER2 expression, has limited effective therapeutic options and unfavorable prognosis. Nicotinamide (NAM), a form of vitamin B3, has shown anti-tumor effects in TNBC, but the underlying regulatory mechanisms remains unclear.

**Methods:** Transcriptomic and proteomic data from TNBC cell lines (BT20, MDA-MB-231, and MDA-MB-468) were analyzed using the ARACNe and VIPER/metaVIPER algorithms to infer regulatory activity and construct gene networks. Differentially expressed genes (DEGs) and proteins (DEPs) were integrated with regulator activity to identify key molecular drivers. Clinical outcomes were assessed using bc-GenExMiner and Kaplan– Meier Plotter, correlation and enrichment analyses were employed ICGC datasets and KEGG pathway mapping.

**Results:** NAM treatment induced distinct but overlapping regulatory activity profiles among TNBC cell lines, with PHGDH, TSPAN1, TACSTD2, and OSBPL6 emerging as shared regulators. Among these, PHGDH, a key enzyme in the serine biosynthesis pathway, showed consistent downregulation across datasets and was associated with poor overall survival (*p*< 0.001). Correlation analysis across three ICGC cohorts identified 384 genes significantly associated with PHGDH, enriched in Glycine, serine and threonine metabolicpathways. Network analysis revealed potential interactions between PHGDH, ERBB3 and other regulatory proteins, suggesting crosstalk between metabolic and signaling pathways.

**Conclusions:** This multi-omics integration highlights PHGDH as a central metabolic regulator linking transcriptional and translational responses to NAM treatment in TNBC. These findings support PHGDH as a potential biomarker and therapeutic target, emphasizing the role of metabolic regulation in TNBC progression and treatment response.

## Introduction

Triple-Negative Breast Cancer (TNBC) is a distinct subtype of breast cancer defined by the absence of estrogen receptor (ER), progesterone receptor (PR), and human epidermal growth factor receptor 2 (HER2) expression. This unique molecular profile limits the effectiveness of hormone- and HER2-targeted therapies, making TNBC particularly challenging to treat.

Accounting for approximately 15% of all breast cancers, TNBC disproportionately affects younger women and is more prevalent among individuals of African and Hispanic descent. Compared with other breast cancer subtypes, TNBC patients face a higher risk of both local and distant recurrence, with metastases more frequently occurring in the brain and lungs rather than bones.If specific markers of TNBC can be identified, it will provide extremely valuable references for its early prediction, diagnosis and timely treatment.

Nicotinamide (NAM), a water-soluble amide form of niacin (vitamin B3), is an important regulator of mitochondrial metabolism and redox reactions. Previous studies have shown that NAM induces cancer cell death in TNBC through mitochondrial dysfunction and reactive oxygen species (ROS) activation, achieved by bifurcating metabolic pathways. However, these studies also revealed variability in gene and protein expression across different TNBC cell lines. Notably, no comprehensive studieson the activity of key regulators or the differential responses among TNBC subgroups are reported.

To address this gap, we analyzed publicly available transcriptomic and proteomic datasets (**Fig. 1A**; **Supplementary Fig. 1**). Specifically, we compared expression profiles across cell lines (**Fig. 1B**), assessed transcription factor activity (**Fig. 1C**), and examined differences in Phosphoglycerate Dehydrogenase(PHGDH) expression among TNBC subtypes (**Fig. 1D**). We further evaluated responses to NAMtreatment and explored associated pathway alterations (**Fig. 1E**).Therefore, we use available data from transcriptomic and proteomic analyses(**Fig. 1A, Supplementary Fig. 1**) to compare differences in expression between cell lines(**Fig. 1B**), assess differences in transcription factor activity(**Fig. 1C**), and analyze differences in PHGDH expression levels across different triple-negative breast cancer subtypes(**Fig. 1D**), as well as changes in response to NAM treatment andalterations in different pathways (**Fig. 1E**).

**Fig. 1.**
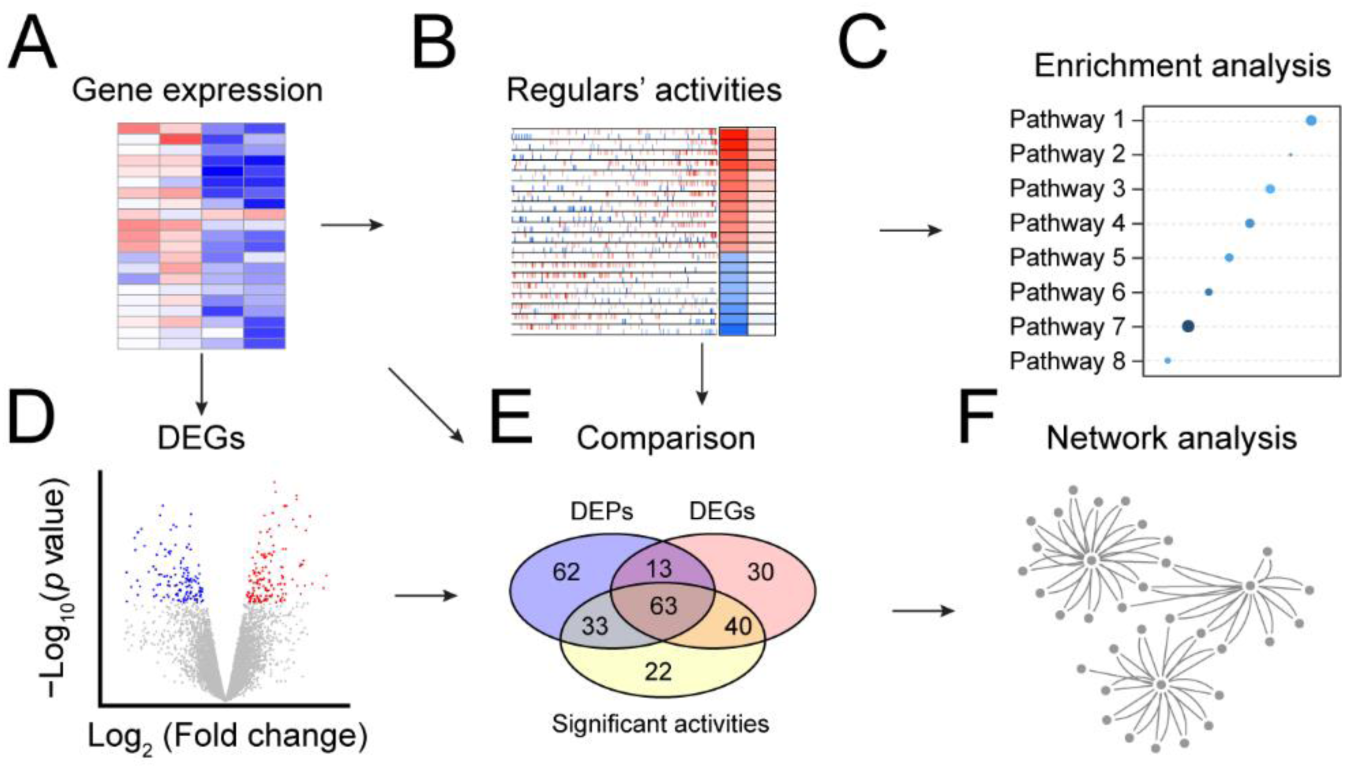
Workflow of the analysis. (A) RNA sequencing data were processed to obtain gene expression profiles across control and treatment samples. (B) Regulatory activities of transcription factors, co-transcription factors, and surface proteins were inferred using the VIPER/metaVIPER algorithm. (C) Functional enrichment analysis was performed to identify significantly enriched KEGG pathways based on active regulators. (D) Differentially expressed genes (DEGs) were identified using the Limma R package. (E) Comparison among differentially expressed proteins (DEPs), DEGs, and significant regulator activities revealed overlapping and unique molecular changes. (F) Network analysis was conducted to visualize interactions among key regulators and target genes, providing an integrated view of transcriptional regulation under treatment conditions.

**Supplementary Fig. 1.**
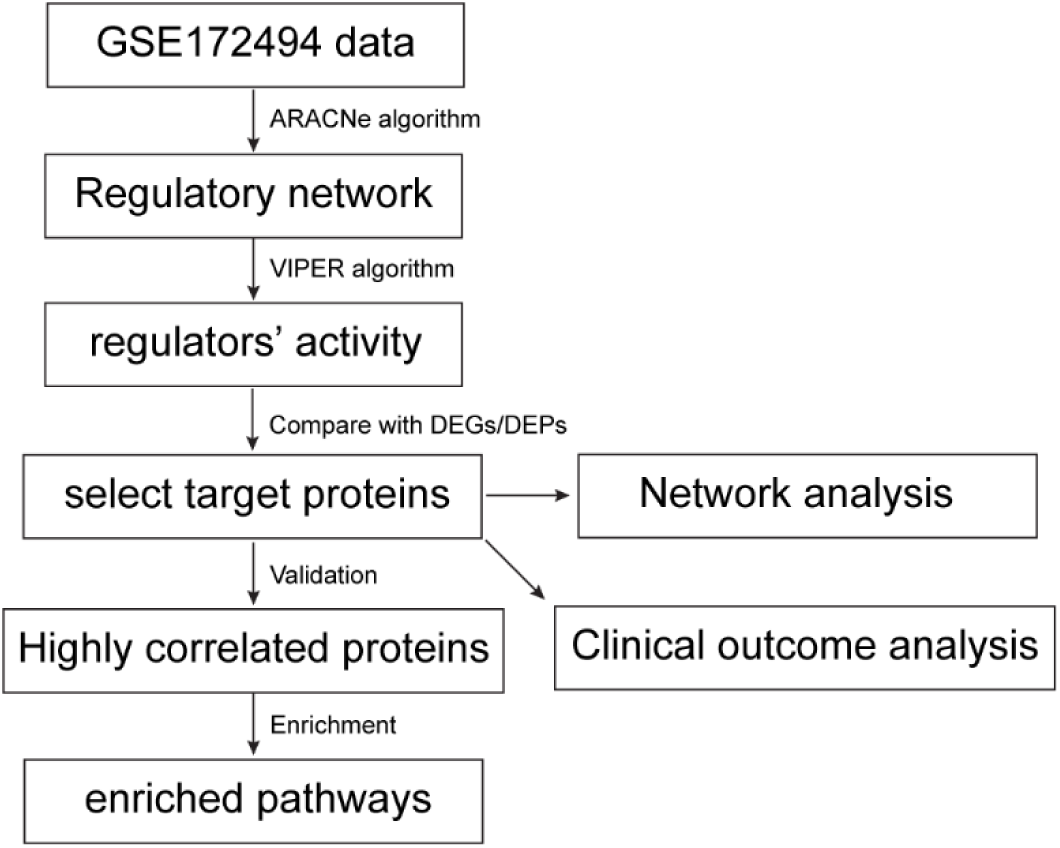
Workflow of the regulators’ activity analysis. RNA sequencing data (GSE172494) were used to build a regulatory network via ARACNe, followed by regulator activity inference using VIPER. Significant regulators were compared with DEGs and DEPs, and selected targets were analyzed through network, clinical outcome, and pathway enrichment analyses.

## Method

### Source of Data

Human breast cancer RNA sequencing data were obtained from the International Cancer Genome Consortium (ICGC) database (https://dcc.icgc.org), including three cohorts from France (BRCA-FR), the United States (BRCA-US), and South Korea (BRCA-KR). Only female breast cancer samples were included in the analysis, and samples with distant metastasis were excluded.

RNA sequencing data for breast cancer cell lines were downloaded from the Gene Expression Omnibus (GEO) database (https://www.ncbi.nlm.nih.gov), encompassing both control and nicotinamide-treated conditions.

### Differentially Expressed Genes (DEG) Analysis

Differential gene expression analysis between cell lines before treatment was performed using the Limma R package^1^. Genes with statistically significant differences in expression were identified as differentially expressed genes (DEGs) based on the predefined thresholds for adjusted p-value and fold change.Regulatory Activity Analysis

Regulatory network construction and activity inference were performed to identify key transcriptional regulators affected by treatment. Using the ARACNe algorithm, and a curated list of 5,542 transcription factors (TFs), co-transcription factors (co-TFs), and surface proteins, a comprehensive interaction network was generated to capture regulatory relationships.

To infer the activity of individual regulators, the VIPER algorithm was applied, which estimates regulator activity from gene expression data through enriched regulon analysis (Alvarez et al., 2016). Furthermore, its extension, metaVIPER, was employed to enhance inference robustness across samples and conditions.

Based on the constructed regulatory networks, the activity of each regulator (TFs, co-TFs, and surface proteins) was calculated using the metaVIPER program. Comparative analyses were then conducted between treated and control samples. Regulators exhibiting statistically significant differences in activity were identified using the Benjamini–Hochberg adjusted p-value threshold.Clinical outcomes analysis

Two online tools were used to analysis the TNBC survival. Kaplan-Meier Plotter (https://kmplot.com/analysis/) was developed with 7,830 samples from 55 independent datasets^2^, included 4,929 patients with breast cancer, and 392 TNBC patients. bc-GenExMiner v5.0 is a statistical mining tool of published annotated breast cancer transcriptomic data (DNA microarrays [n = 11,552] and RNA-seq [n = 5,023]). ^3^ It offers the possibility to explore gene-expression of genes of interest in breast cancer. There is, 1015 TNBC patients with subtype of BLIA.

### Correlation analysis

Pearson correlation analysis was employed using R tools to assess the relationships between genes and other expression levels. This statistical *p* value is 0.05.

### Enrichment analysis

Enrichment analysis was employed using “RichR” R package to explore Kyoto Encyclopedia of Genes and Genomes (KEGG) pathways. This statistical *p* value is 0.05.

### Network analysis

The relationship of genes was explore using online tool of STRING 12.0. The net was generated by Cytoscape.

## Results

### 2.1 Regulators’ activity analysis after treatment

To set up a regulatory network between transcription factors, surface proteins, co-transcription factors, and genes, RNA sequencing data from BT20, MDA-MB-231, and MDA-MB-468 cell lines in the GSE172494 dataset ^4^ were analyzed. After data preprocessing, including filtering and normalization, 26,266 genes were subjected to ARACNe analysis. Using a curated list of 5,542 regulators, a comprehensive network comprising 1,223,258 interactions was constructed.To compare treatment and control conditions and explore the effects of nicotinamide, the VIPER algorithm was applied to identify master regulators. In MDA-MB-468 cells, 169 upregulated and 255 downregulated regulators were detected, whereas MDA-MB-231 cells exhibited 73 upregulated and 76 downregulated regulators, and BT20 cells showed 129 upregulated and 192 downregulated regulators. Comparative analysis across cell lines revealed 73 shared regulators between MDA-MB-468 and BT20, and 15 common regulators across all three cell lines (**Fig. 2A, Supplementary Table 1**).To explore functional implications of the significant regulators, KEGG pathway enrichment analysis was performed (**Fig. 2B**). Notably, two pathways—Pathways in cancer and Leukocyte transendothelial migration—were shared across all three cell lines (**Supplementary Table 2**). In MDA-MB-231, the most significantly enriched pathways included Neuroactive ligand–receptor interaction, Renal cell carcinoma, and Chemokine signaling pathway, with three unique pathways—Amyotrophic lateral sclerosis, Neuroactive ligand–receptor interaction, and Renal cell carcinoma (**Fig. 2B**). MDA-MB-468 cells showed enrichment in the B cell receptor signaling pathway, Wnt signaling pathway, and Melanogenesis (**Fig. 2C**). In BT20, the most significant pathways comprised Leukocyte transendothelial migration, Fc epsilon RI signaling pathway, and Colorectal cancer (**Fig. 2D**).

**Fig. 2.**
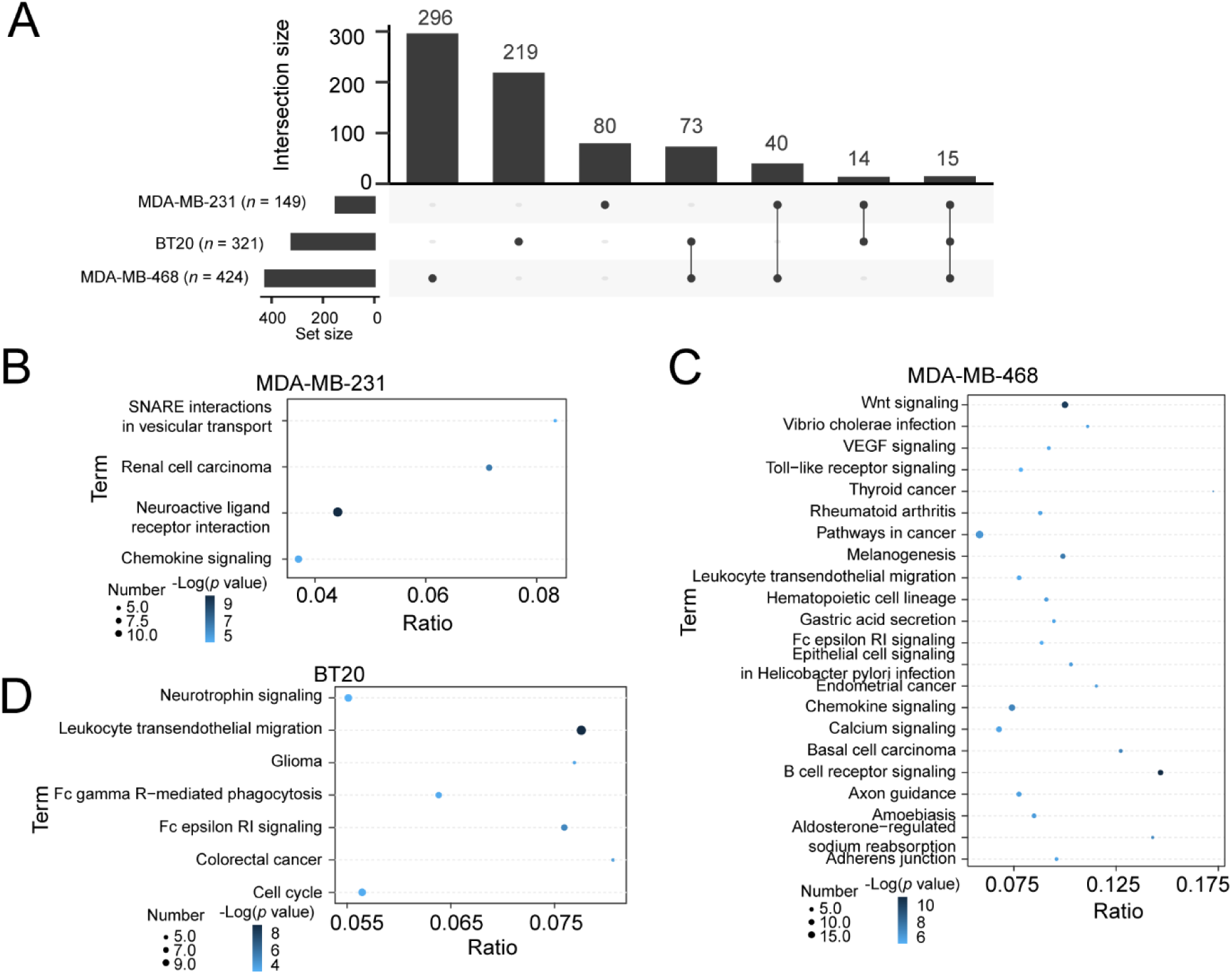
Regulators’ activity analysis between treatment and control. (A) Overlap of significantly altered regulators among MDA-MB-231, MDA-MB-468, and BT20 cell lines. (B–D) KEGG pathway enrichment analysis of regulators with significant activity changes in MDA-MB-231 (B), MDA-MB-468 (C), and BT20 (D) cells. The dot size represents the number of genes enriched in each pathway, while the dot color corresponds to the statistical significance level (−log₁₀p-value), with darker colors indicating higher significance. Pathways shared among cell lines included Pathways in cancer and Leukocyte transendothelial migration.

To summarize the significant results, different TNBC cell subtypes exhibited distinct regulatory activity patterns, with MDA-MB-468 and BT20 showing more similar trends compared to MDA-MB-231. When comparing baseline and post-treatment conditions, 73 regulators were shared between MDA-MB-468 and BT20, of which 68 (93%) displayed consistent trends—47 downregulated and 21 upregulated (**Supplementary Table 1**). Between MDA-MB-231 and MDA-MB-468, 40 significantly altered regulators were shared, with 29 (72.5%) showing the same directional change—11 downregulated and 18 upregulated. In contrast, comparing MDA-MB-231 and BT20, 14 significantly altered regulators were identified, with 12 (86%) exhibiting concordant trends—7 downregulated and 5 upregulated. To compare the regulator activities with transcriptomic and proteomic changes, we next examined the relationships among regulatory activities, differentially expressed genes (DEGs), and differentially expressed proteins (DEPs) after treatment. A total of 73 regulators were shared between MDA-MB-468 and BT20, among which 43 corresponding DEGs were identified, and 37 genes exhibited consistent trends in activity and expression (**Fig. 3C**). Before treatment, a marked difference was noted between MDA-MB-468 and BT20, with six overlapping proteins, four of which maintained the same direction of change after treatment.

**Fig. 3.**
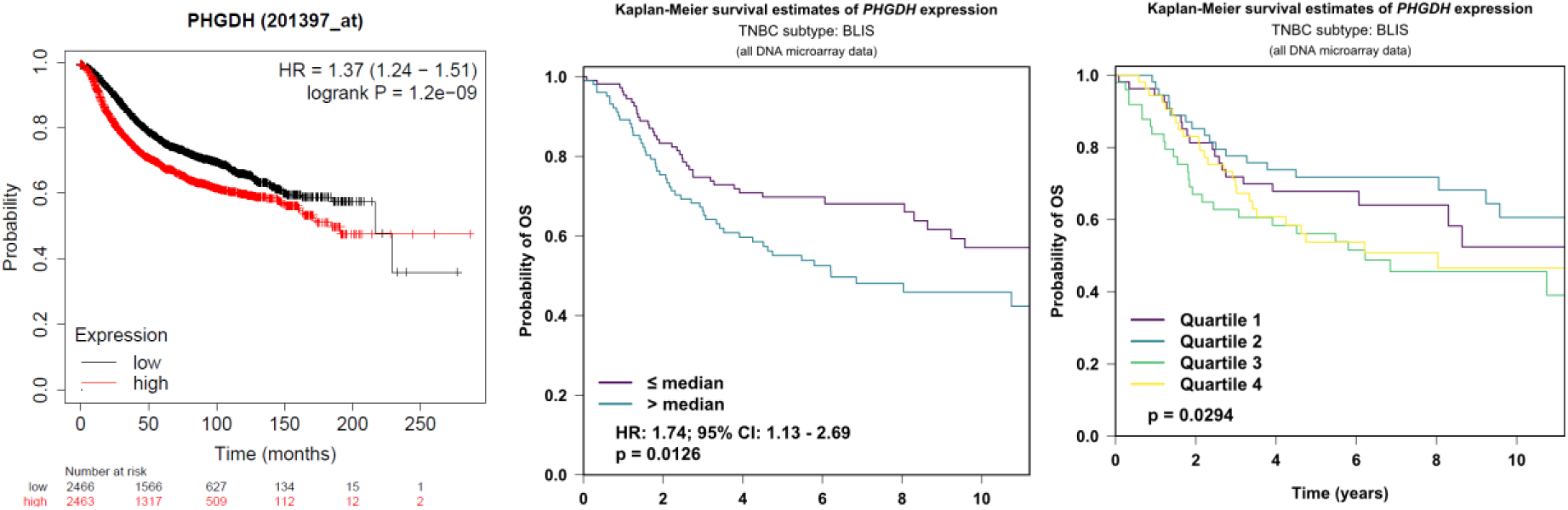
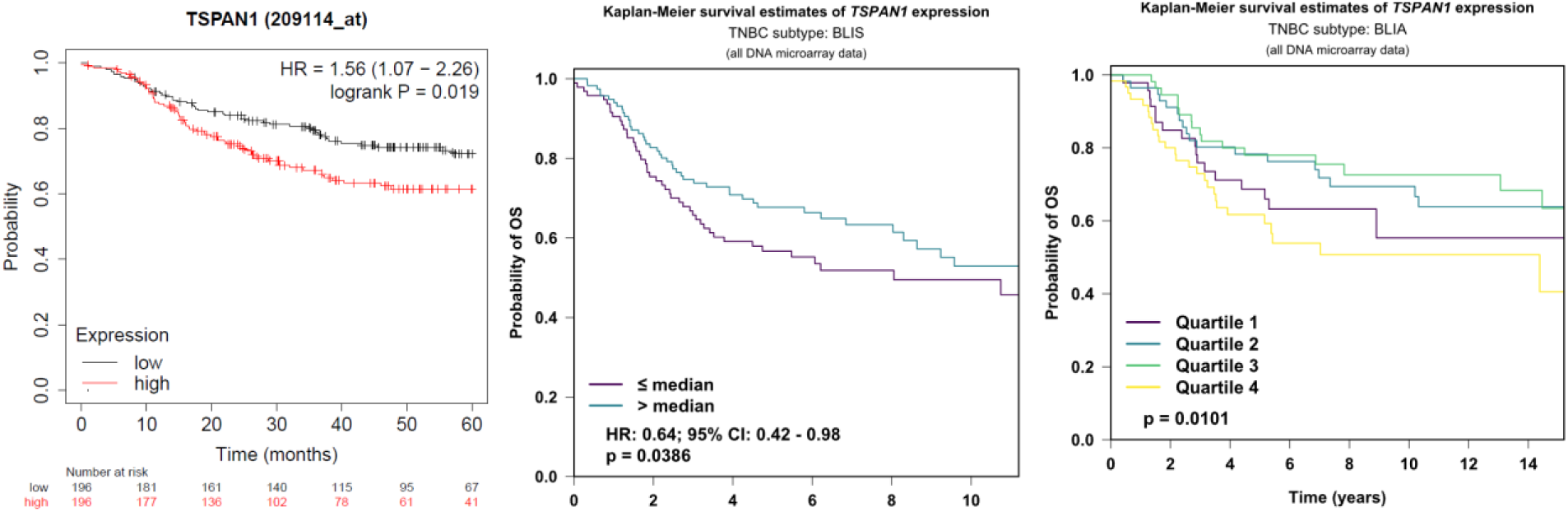
Survival curves from two online analysis tools. Kaplan–Meier survival analyses of PHGDH (top) and TSPAN1 (bottom) expression in breast cancer patients. The left panels show results from bc-GenExMiner, comparing high and low expression groups with corresponding hazard ratios (HR) and log-rank *p*-values. The middle and right panels present analyses from the Kaplan–Meier Plotter, where patients were divided by median (middle) or quartile (right) expression levels. Higher PHGDH expression was associated with poorer overall survival, whereas TSPAN1 showed inconsistent effects across datasets.

Importantly, **four genes—TSPAN1, TACSTD2, PHGDH, and OSBPL6—**showed consistent trends across regulatory activity, gene expression, and protein abundance following treatment. These genes also displayed significant differences when compared with MDA-MB-231, both in expression levels and inferred activities (**Supplementary Table S7**), highlighting their potential roles as key mediators of nicotinamide-induced effects in TNBC cells.To evaluate the clinical relevance of the identified regulators, Cox proportional hazards regression analysis was performed using two online tools: bc-GenExMiner and Kaplan–Meier Plotter.

In bc-GenExMiner, higher expression of PHGDH was identified as a significant risk factor for overall survival (OS) in breast cancer patients (n = 1,879). Patients with low PHGDH expression had a median OS of 130 months, which was significantly longer than that of patients with high expression (68 months, *p* = 2.0 × 10⁻⁸; **Fig. 3A**). Consistent findings were obtained using the Kaplan–Meier Plotter, where patients were dichotomized by median expression. High PHGDH expression was again associated with poorer prognosis, with a hazard ratio (HR) of 1.74 (95% CI: 1.13–2.69, *p* = 0.01; **Fig. 3B**). When analyzed by quartiles, this association remained significant (*p* = 0.03; **Fig. 3C**).

In contrast, the results for TSPAN1 were inconsistent across analyses. In bc-GenExMiner, higher TSPAN1 expression was associated with worse disease-free survival (DFS) in breast cancer patients. The median OS was 108 months for patients with low expression versus 85 months for those with high expression (*p* = 0.02; **Fig. 3D**). However, in the Kaplan–Meier Plotter, when patients were divided by median expression, higher TSPAN1 expression appeared to be a protective factor, with an HR of 0.64 (95% CI: 0.42–0.98, *p* = 0.04; **Fig. 3E**). When analyzed by quartiles, the association remained significant (*p* = 0.01; **Fig. 3F**).

PHGDH, a gene highly expressed in TNBC cell lines and tissues, has been highlighted in multiple studies^5–8^. As an enzyme catalyzing the first committed step of the serine biosynthesis pathway, PHGDH converts 3-phosphoglycerate to 3-phosphohydroxypyruvate, a crucial step in serine production. Elevated PHGDH expression has been observed across multiple cancer types, particularly in Triple-Negative Breast Cancer (TNBC), where it contributes to tumor growth and metabolic adaptation. Due to its central role in cancer metabolism, PHGDH is increasingly recognized as a promising therapeutic target for TNBC.

### 2.4 Correlation Analysis in Human RNA Sequencing

To validate the biological functions of PHGDH, RNA sequencing data from three breast cancer cohorts were obtained from the International Cancer Genome Consortium Accelerating Research in Genomic Oncology (ICGC ARGO) database (https://docs.icgc-argo.org/).

In the BRCA-FR cohort, after data filtering and normalization, 20,039 genes from 95 samples were included in the correlation analysis with PHGDH expression. Using a significance cutoff of p < 0.01, 2,635 genes showed a significant correlation with PHGDH. In the BRCA-KR cohort, after similar preprocessing, 14,859 genes from 47 samples were analyzed, revealing 478 genes significantly correlated with PHGDH (p < 0.01). In the BRCA-US cohort, 17,814 genes from 524 samples were examined, and 11,431 genes were significantly correlated with PHGDH (p < 0.01), among which 300 genes had an absolute correlation coefficient greater than 0.5.

Across the three cohorts, 384 genes were consistently correlated with PHGDH. Functional enrichment analysis revealed that these genes were predominantly involved in Glycine, serine and threonine metabolism and Metabolic pathways (**Supplementary Fig. 2A**), supporting the central role of PHGDH in cellular metabolic regulation.

**Supplementary Fig. 2.**
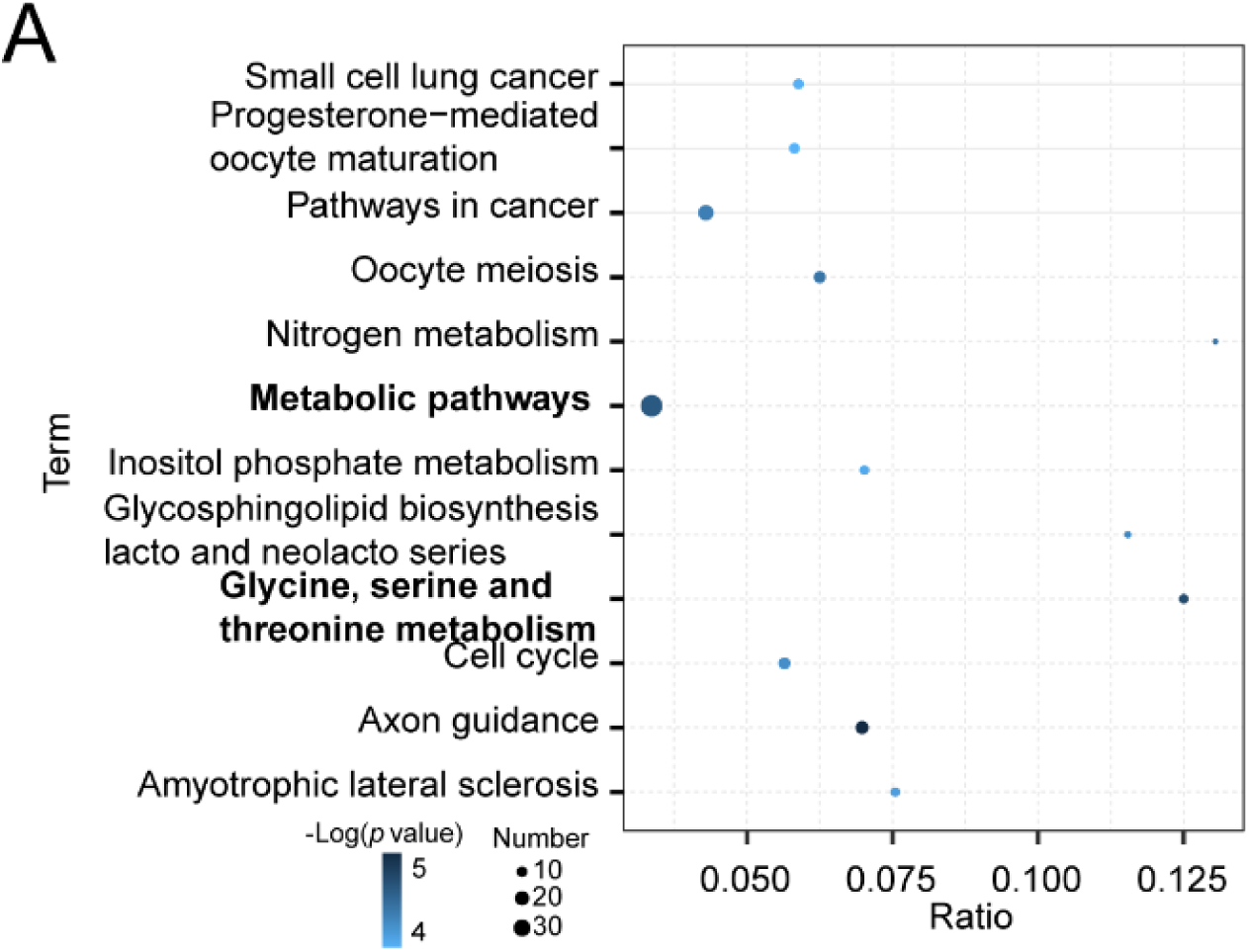
Enrichment analysis by the PGHDH highly correlated genes. KEGG pathway enrichment analysis of genes significantly correlated with PHGDH across three ICGC breast cancer cohorts. The dot size represents the number of genes enriched in each pathway, and the dot color indicates the statistical significance level (−log₁₀ p-value). Prominent pathways include Metabolic pathways and Glycine, serine, and threonine metabolism, highlighting PHGDH’s central role in cellular metabolism.

### 2.5 Network Analysis of PHGDH

To further investigate the regulatory landscape associated with PHGDH, a gene interaction network was constructed using the ARACNe algorithm. The resulting network contained 903 genes directly connected to PHGDH. After filtering for regulators exhibiting significant activity changes, the refined network was visualized in Supplementary Fig. 3.

**Supplementary Fig. 3.**
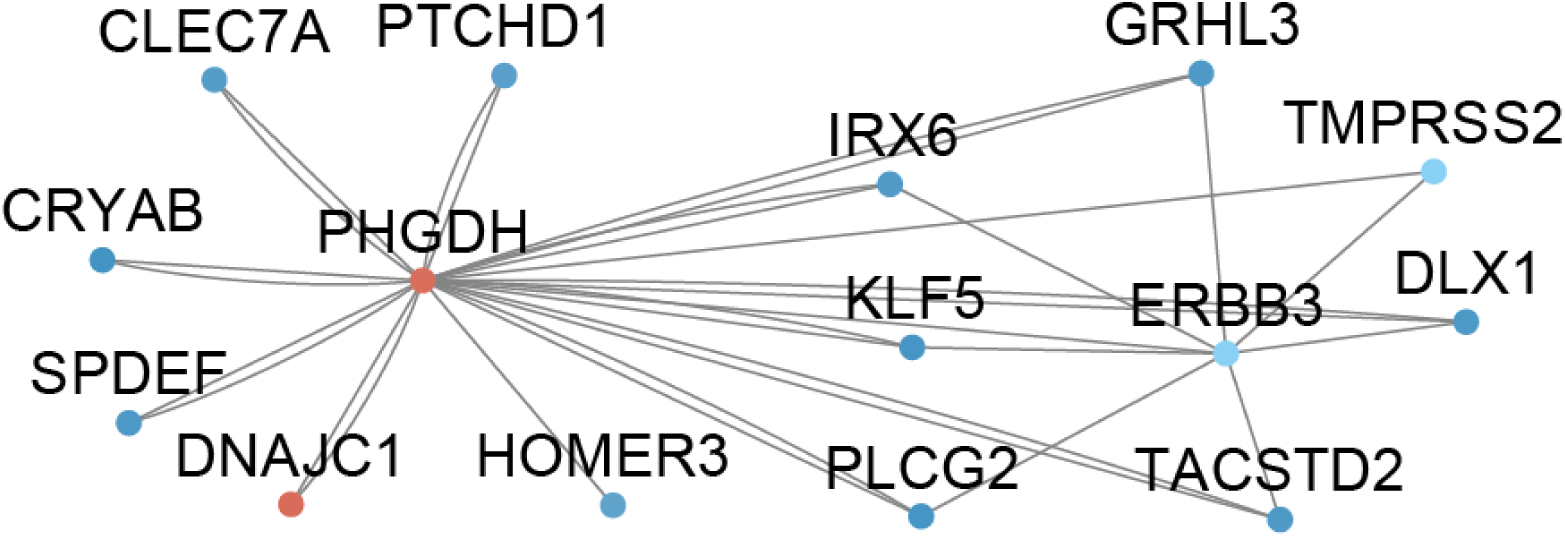
Network with PHGDH. Regulatory network centered on PHGDH constructed using the ARACNe algorithm. Nodes represent interacting genes, and edges indicate predicted regulatory connections. Key interacting proteins include DNAJC1, ERBB3, and TACSTD2.

Within this network, 14 proteins were identified as directly interacting with PHGDH, suggesting potential co-regulatory or functional associations. Additionally, seven regulators were found to interact with ERBB3, indicating possible crosstalk between PHGDH-associated metabolic regulation and ERBB3-mediated signaling pathways.

## Discussion

Using the ARACNe algorithm, we established a regulatory network that predicts the activity of key regulators and identifies their connections with downstream genes and proteins. This approach enabled the systematic analysis of transcription factors, co-factors, and surface proteins in nicotinamide-treated TNBC cell lines. By integrating VIPER-based activity inference with gene expression and protein abundance data, we identified PHGDH, TSPAN1, TACSTD2, and OSBPL6 as central regulators consistently altered across multiple cell lines. Among them, PHGDH showed strong metabolic pathway enrichment and was significantly associated with poor clinical outcomes, highlighting its potential role as a metabolic driver and therapeutic target in TNBC.The ARACNe algorithm generates a list of interacting probe set pairs ranked by Mutual Information (MI) values and associated p-values, where a higher MI indicates stronger statistical dependence between genes ^9^. Biologically, an interaction between gene A and gene B suggests that both participate in the same physiological process; moreover, if gene A is a transcription factor (TF) and gene B is a non-TF, this implies that gene A may act as a transcriptional regulator of gene B.

Previous studies have primarily examined the effects of nicotinamide (NAM) on gene and protein expression levels. In contrast, this study focused on regulatory activity, which bridges transcriptional regulation and translational output. By investigating how transcription factors and their targets respond to NAM treatment, we link RNA-level regulation with protein-level outcomes, offering insights into the mechanisms driving cellular adaptation and dysfunction. Such regulatory insights are critical, as disruptions in RNA processing or protein modification have been implicated in various cancers, highlighting the potential of targeting these pathways for therapeutic intervention.

### Differential responses with NAM in TNBC

Interestingly, our findings align with previous studies that reported differential responses among the BT20, MDA-MB-231, and MDA-MB-468 cell lines, yet we observed similar trends in regulatory activities across these lines. This suggests the presence of shared regulatory pathways that might underlie common adaptive mechanisms, despite their distinct molecular profiles and phenotypic behaviors. Such commonalities could provide valuable insights into therapeutic strategies, as targeting these shared regulators may yield broader effects across different breast cancer subtypes. Furthermore, this convergence on similar regulatory activities underscores the plasticity of cancer cells, highlighting potential biomarkers for monitoring treatment responses and disease progression in breast cancer.

### The Oncogenic Role of PHGDH in TNBC: Mechanisms and Therapeutic Implications

Our data corroborate previous findings that PHGDH, the first and rate-limiting enzyme in the serine biosynthesis pathway (SSP), is upregulated in aggressive TNBC models and is susceptible to metabolic perturbation, as evidenced by its consistent downregulation following NAM treatment in both BT-20 and MDA-MB-468 cells. While the initial discovery of PHGDH gene amplification highlighted its importance, our understanding of its multifaceted oncogenic role has significantly expanded. This discussion will contextualize our findings within the broader framework of how PHGDH regulates TNBC biology and why it represents a promising but complex therapeutic node.

### Beyond Serine Production: The Multifaceted Mechanisms of PHGDH-Driven Tumorigenesis

The fundamental role of PHGDH in converting 3-phosphoglycerate into 3-phosphohydroxypyruvate is to fuel the de novo synthesis of serine and glycine. This function is undeniably crucial for satisfying the anabolic demands of rapidly proliferating cancer cells. The resulting serine is essential for nucleotide synthesis, lipid biosynthesis, and the maintenance of redox balance through glutathione production. However, to view PHGDH solely as a supplier of amino acids represents a significant oversimplification of its function. Its oncogenic influence extends into a sophisticated network of signaling and epigenetic regulation^10^.

Our observation that PHGDH is uniquely sensitive to NAM—a precursor for NAD+ and an inhibitor of NAD+-consuming enzymes like sirtuins—suggests a deeper layer of regulation that may involve mitochondrial function and energy sensing^11^. The interplay between NAD+ metabolism and PHGDH activity suggests a potential feedback mechanism where cellular energy status fine-tunes anabolic flux. Furthermore, PHGDH-derived metabolites are potent signaling molecules. For instance, serine availability is a key activator of the mTORC1 pathway, a master regulator of cell growth^12^. Perhaps most intriguingly, the PHGDH is essential for mounting a type I interferon (IFN-β) response, promoting the mitochondrial antiviral signaling pathway and facilitating IRF3 nuclear translocation^13^. This positions PHGDH not only as a direct anabolic driver but also as a key regulator of the tumor immune microenvironment.

### PHGDH is a Viable Therapeutic Target in TNBC: From Concept to Clinic

The aggressive nature of TNBC, characterized by a lack of actionable targets for endocrine or HER2-directed therapy, creates an urgent need for novel treatment strategies. Targeting metabolic vulnerabilities, such as dependence on the SSP, offers a promising avenue. Our data, showing that two distinct TNBC cell lines (BT-20 and MDA-MB-468) respond to a lower dose of NAM with concomitant PHGDH downregulation, provide preliminary support for the pharmacological vulnerability of this pathway.Moreover, the fact that both responsive cell types (BT-20 and MDA-MB-468) exhibited this effect at a lower dose of NAM further underscores this vulnerability ^4^.

The development of specific small-molecule PHGDH inhibitors such as CBR-5884, NCT-502^14^has putthis concept from theory toward practice. Preclinical studies demonstrate that PHGDH inhibition selectively impairs the growth and viability of PHGDH-dependent TNBC cells, validating its role as a cancer fitness gene. Importantly, the therapeutic potential may extend beyond monotherapy. PHGDH inhibition can create synthetic lethal vulnerabilities. For example, by depleting nucleotide pools, it can synergize with DNA-damaging agents like chemotherapeutics or PARP inhibitors^15,16^. Combining PHGDH inhibitors with a low serine/glycine diet could exacerbate metabolic stress in cancer cells while sparing normal tissues^17^. Our work suggests that exploring combinations with NAD+-modulating agents like NAM could be another fruitful area of investigation, potentially targeting cancer metabolism from multiple angles.

### Future Directions and Translational Challenges

Despite the compelling rationale, several challenges must be addressed to successfully translate PHGDH targeting into the clinic. First, patient stratification is paramount. Not all TNBCs are PHGDH-addicted; thus, robust biomarkers^18^—such as PHGDH protein expression by immunohistochemistry or gene amplification status—must be developed to identify the patient population most likely to benefit.

Second, the issue of potential on-target toxicity must be carefully evaluated. Given the high expression of PHGDH in the brain and its role in neurological function^19^, inhibiting systemic PHGDH could have adverse effects. Strategies to mitigate this, such as intermittent dosing or leveraging the blood-brain barrier, will be crucial^20^. Finally, as with any targeted therapy, resistance mechanisms are likely to emerge. Cancer cells may upregulate serine transporters to scavenge exogenous serine or rewire their metabolic networks to bypass the blockade^5^. Understanding these adaptive responses will be key to developing effective combination regimens that deliver durable responses.

## Conclusion

In conclusion, our integrated multi-omics analysis revealed that a subset of TNBC models (namely MDA-MB-468 and BT-20) share common regulatory vulnerabilities to NAM treatment, with the serine biosynthetic enzyme PHGDH emerging as a consistently downregulated core node. This finding positions PHGDH at a critical nexus, integrating metabolic flux, transcriptional regulation, and signals from the tumor microenvironment to drive TNBC progression. It is, therefore, more than a metabolic enzyme; it is a central regulator of oncogenic metabolism. While translational hurdles remain, the continued development of potent and specific inhibitors, coupled with a deeper understanding of its biology and reliable biomarkers, positions PHGDH as a leading candidate for a new class of metabolic therapies aimed at overcoming the therapeutic challenges of TNBC.

## Data Availability

All data produced in the present study are available upon reasonable request to the authors

## Authors’ contributions

Yijun Xie* and Ziyu Jiang* contributed equally to this work. Conceptualization and design: Y.X., Z.J., H.L., L.H.; Data analysis and interpretation: Y.X., Z.J., Z.H., Z.C.; Literature search and original draft preparation: Y.X., Z.J., H.L., L.H.; Manuscript editing and supervision: Z.H., Z.C., H.L., L.H. All authors reviewed and approved the final manuscript.

## Acknowledgements

Not applicable.

## Ethics approval and consent to participate

Not applicable.

## Consent for publication

Not applicable.

## Availability of data and materials

The transcriptomic and proteomic data analyzed in this study were derived from the Triple-Negative Breast Cancer (TNBC) cell lines BT20, MDA-MB-231, and MDA-MB-468. The analysis pipelines included the ARACNe and VIPER/metaVIPER algorithms. Clinical correlation analyses were performed using the public databases bc-GenExMiner, Kaplan-Meier Plotter, and International Cancer Genome Consortium (ICGC) datasets. Pathway enrichment analysis was conducted using KEGG. All datasets utilized are publicly available as referenced in the manuscript.

## References

1. Ritchie ME, Phipson B, Di Wu, et al. limma powers differential expression analyses for RNA-sequencing and microarray studies. Nucleic acids research. 2015;43(7):e47.

2. Győrffy B. Survival analysis across the entire transcriptome identifies biomarkers with the highest prognostic power in breast cancer. Computational and structural biotechnology journal. 2021;19:4101–4109.

3. Jézéquel P, Campone M, Gouraud W, et al. bc-GenExMiner: An easy-to-use online platform for gene prognostic analyses in breast cancer. Breast cancer research and treatment. 2012;131(3):765–775.

4. Jung M, Lee K-M, Im Y, et al. Nicotinamide (niacin) supplement increases lipid metabolism and ROS-induced energy disruption in triple-negative breast cancer: Potential for drug repositioning as an anti-tumor agent. Molecular oncology. 2022;16(9):1795–1815.

5. Kim SK, Jung WH, Koo JS. Differential expression of enzymes associated with serine/glycine metabolism in different breast cancer subtypes. PloS one. 2014;9(6):e101004.

6. Metcalf S, Dougherty S, Kruer T, et al. Selective loss of phosphoserine aminotransferase 1 (PSAT1) suppresses migration, invasion, and experimental metastasis in triple negative breast cancer. Clinical & experimental metastasis. 2020;37(1):187–197.

7. Gromova I, Gromov P, Honma N, et al. High level PHGDH expression in breast is predominantly associated with keratin 5-positive cell lineage independently of malignancy. Molecular oncology. 2015;9(8):1636–1654.

8. Zhang X, Bai W. Repression of phosphoglycerate dehydrogenase sensitizes triple-negative breast cancer to doxorubicin. Cancer chemotherapy and pharmacology. 2016;78(3):655–659.

9. Chávez Montes RA, Coello G, González-Aguilera KL, Marsch-Martínez N, Folter S de, Alvarez-Buylla ER. ARACNe-based inference, using curated microarray data, of Arabidopsis thaliana root transcriptional regulatory networks. BMC plant biology. 2014;14:97.

10. Cheng B, Peng P, Chen S, et al. Phosphoglycerate dehydrogenase stabilizes protein kinase C delta type mRNA to promote hepatocellular carcinoma progression. Signal transduction and targeted therapy. 2025;10(1):236.

11. Barnabas GD, Lee JS, Shami T, et al. Serine Biosynthesis Is a Metabolic Vulnerability in IDH2-Driven Breast Cancer Progression. Cancer research. 2021;81(6):1443–1456.

12. Rinaldi G, Pranzini E, van Elsen J, et al. In Vivo Evidence for Serine Biosynthesis-Defined Sensitivity of Lung Metastasis, but Not of Primary Breast Tumors, to mTORC1 Inhibition. Molecular cell. 2021;81(2):386–397.e7.

13. Li X, Huang Y, Liu X, et al. Classical swine fever virus inhibits serine metabolism-mediated antiviral immunity by deacetylating modified PHGDH. mBio. 2024;15(10):e0209724.

14. Islam MM, Kasana S, Priya S, Kurmi BD, Gupta GD, Patel P. Harnessing PHGDH Inhibition for Cancer Therapy: Mechanisms, SAR, Computational Aspects, and Clinical Potential. Archiv der Pharmazie. 2025;358(8):e70083.

15. Zhang X, Sun M, Jiao Y, Lin B, Yang Q. PHGDH Inhibitor CBR-5884 Inhibits Epithelial Ovarian Cancer Progression via ROS/Wnt/β-Catenin Pathway and Plays a Synergistic Role with PARP Inhibitor Olaparib. Oxidative medicine and cellular longevity. 2022;2022:9029544.

16. van Nyen T, Planque M, van Wagensveld L, et al. Serine metabolism remodeling after platinum-based chemotherapy identifies vulnerabilities in a subgroup of resistant ovarian cancers. Nature communications. 2022;13(1):4578.

17. Muthusamy T, Cordes T, Handzlik MK, et al. Serine restriction alters sphingolipid diversity to constrain tumour growth. Nature. 2020;586(7831):790–795.

18. van Wagensveld L, van Nyen T, Annibali D, et al. High expression of phosphoglycerate dehydrogenase predicts poor outcome in patients with high-grade serous ovarian cancer. The oncologist. 2024;29(9):e1231–e1234.

19. Kinoshita MO, Shinoda Y, Sakai K, et al. Selective upregulation of 3-phosphoglycerate dehydrogenase (Phgdh) expression in adult subventricular zone neurogenic niche. Neuroscience letters. 2009;453(1):21–26.

20. Cali Daylan AE, Leone JP. Targeted Therapies for Breast Cancer Brain Metastases. Clinical breast cancer. 2021;21(4):263–270.

